# Intrinsic functional connectivity of right dorsolateral prefrontal cortex and hippocampus subregions relates to emotional and sensory-perceptual properties of intrusive trauma memories

**DOI:** 10.1101/2024.10.12.24314977

**Authors:** Quentin Devignes, Kevin J. Clancy, Boyu Ren, Yara Pollmann, Justin T. Baker, Isabelle M. Rosso

## Abstract

**Background:** Trauma-related intrusive memories (TR-IMs), unwanted and vivid, are core symptoms of posttraumatic stress disorder (PTSD). Prior research links voluntary TR-IM suppression to inhibitory control of the right dorsolateral prefrontal cortex (dlPFC) over the hippocampus (HPC). However, the potential relevance of tonic resting-state inhibition has not been examined, nor has the functional differentiation of the anterior and posterior hippocampus (aHPC/HPC). This study examined relationships of TR-IM frequency and properties with resting-state negative coupling between the right dlPFC and right aHPC/pHPC in trauma-exposed individuals with PTSD symptoms.

**Methods:** Participants (N=109; 88 female) completed two weeks of ecological momentary assessments capturing TR-IM frequency and properties (intrusiveness, emotional intensity, vividness, visual properties, and reliving). Using 3T resting-state magnetic resonance imaging, participant-specific 4-mm spheres were placed at the right dlPFC voxel most anticorrelated with the right aHPC and pHPC. Quasi-Poisson and linear mixed-effects models assessed relationships of TR-IM frequency and properties with right dlPFC-right aHPC and pHPC anticorrelation.

**Results:** TR-IM emotional intensity was positively associated with right dlPFC-aHPC connectivity, while vividness and visual properties were linked to right dlPFC-pHPC connectivity. No significant associations were found between TR-IM frequency, intrusiveness, or reliving, and anticorrelation with either HPC subregion.

**Conclusions:** This study provides novel insights into the neural correlates of TR-IMs, highlighting the relevance of intrinsic negative coupling between the right dlPFC and aHPC/pHPC to their emotional impact and perceptual properties. Further research on inhibitory mechanisms in this circuit could improve understanding of component processes of intrusive reexperiencing, a severe and treatment-refractory PTSD symptom.

## INTRODUCTION

Trauma-related intrusive memories (TR-IMs) are involuntary recollections of a traumatic event and core symptoms of posttraumatic stress disorder (PTSD) (1,2). These memories are typically vivid, sensory-based (predominantly visual) experiences that are accompanied by intense negative emotions and a sense of reliving the trauma in the present (3). TR-IMs often disrupt daily functioning (4–6) and predict the development and severity of other PTSD symptoms (4,7–9). Despite their clinical significance, the neurophysiological bases and phenomenological properties of TR-IMs remain unclear. This may be partly due to historical reliance on retrospective diagnostic assessments, which focus mostly on the frequency and intensity of TR-IMs but minimally on their phenomenological qualities (1,10). Therefore, there is a need for investigations combining neuroimaging with detailed assessments of TR-IM frequency and properties to identify brain targets for interventions aimed at mitigating their occurrence and impact.

Functional neuroimaging research has highlighted the critical role of the right dorsolateral prefrontal cortex (dlPFC) in voluntarily suppressing intrusive memories by inhibiting hippocampus-mediated retrieval (11–15). Studies have demonstrated that when individuals are instructed to suppress intrusive recollections, right dlPFC activity increases while right hippocampus (HPC) activity decreases, with increased negative functional coupling between the two regions (13–18). However, individuals with PTSD often fail to show this expected increase in right dlPFC-right HPC negative coupling during memory suppression attempts, which correlates with more severe intrusion symptoms and the persistence of intrusive memories over time (13,19–21). In healthy adults, volitional suppression attempts engaging this top-down inhibition of HPC activity lead to a gradual and sustained reduction in the frequency, vividness, and emotional intensity of intrusive memories over time (14,15). This suggests the strengthening of a tonic inhibitory mechanism that weakens memories in the longer term, diminishing both their recurrence and phenomenological features (22,23). Although the neural mechanisms underlying this sustained inhibition are underexplored, research suggests it involves a decline in the reactivation of memory-related regions, particularly the HPC (23). Thus, individual differences in the intrinsic resting-state negative coupling of right dlPFC-HPC pathways may support the tonic control of unwanted memory retrieval and contribute to TR-IM frequency and properties in PTSD. In addition, because resting-state connectivity patterns have been shown to predict task-based activity and cognitive performance (24,25), the intrinsic coupling of the right dlPFC-HPC circuit may have critical implications for its effective engagement during TR-IM suppression.

An important consideration when studying HPC function is the partially distinct roles of its anterior and posterior divisions during autobiographical memory retrieval. The anterior (aHPC) and posterior (pHPC) HPC exhibit different structural and functional connectivity patterns with other brain regions (26–29) and are implicated in different component processes of memory retrieval (30–37). The aHPC is primarily involved in the initial construction of memories, facilitating the retrieval of central themes or the gist, along with the emotional context (30–39). In contrast, the pHPC is more engaged during the elaboration phase, retrieving fine-grained details, including the memory’s sensory-perceptual qualities (30–37). As this process unfolds, dlPFC connectivity with both aHPC and pHPC allows modulation of these retrieval phases, enabling a coherent reinstatement of mnemonic content into consciousness (14,15). However, most studies examining neural correlates of intrusive memory suppression have not distinguished between hippocampal subregions (14,18,23,40–44), although a few reported that aHPC activity was more correlated with intrusion frequency (13,15). Furthermore, no studies have investigated whether the tonic, resting-state functioning of the dlPFC, aHPC, and pHPC are related to intrusive memory retrieval.

In a recent investigation by our group, we sought to characterize relationships between functional and structural features of a/pHPC networks and TR-IM frequency and properties in adults with PTSD. Our first report on resting-state data used whole-brain coactivation pattern analyses with the aHPC and pHPC as seeds. This revealed partially distinct associations of aHPC/pHPC positive and negative resting-state functional connectivity with TR-IM properties and activity in other brain regions, including the prefrontal cortex (45). These findings underscore the importance of considering aHPC and pHPC separately when investigating TR-IM neurophysiology (45). However, this approach did not specifically examine the negative functional coupling between the right dlPFC-right a/pHPC (45), a circuit that, as noted earlier, may play a critical inhibitory role in suppressing intrusive memories. Furthermore, the identified a/pHPC coactivation patterns were not associated with TR-IM frequency (45), suggesting the involvement of a different or specific circuit in the occurrence of TR-IMs. In support of this, we recently demonstrated that more frequent occurrence of TR-IMs was significantly associated with lower structural synchronization between the aHPC and other brain regions involved in autobiographical memory, including the dlPFC (46). Collectively, these findings underscore the pertinence of subdividing the HPC into anterior and posterior subregions to deepen our understanding of the neural correlates of TR-IMs.

In summary, understanding the mechanisms of TR-IM frequency and properties may be advanced by examining right dlPFC-right HPC resting-state negative coupling while considering the functional differentiation between the aHPC and pHPC. This study leveraged data from trauma-exposed adults who completed ecological momentary assessment (EMA) of TR-IM frequency and properties, along with resting-state functional MRI (rsfMRI). Our primary hypothesis was that weaker right dlPFC-right HPC resting-state negative coupling, reflecting diminished tonic modulation, would be associated with both TR-IM frequency and properties.

Specifically, building on prior research implicating the aHPC and pHPC in distinct aspects of autobiographical memory, we hypothesized that: (a) higher frequency, emotional intensity, and intrusiveness of TR-IMs would be significantly associated with weaker negative coupling between right dlPFC and right aHPC, but not pHPC; and (b) higher vividness, enhanced visual properties, and a stronger sense of reliving of TR-IMs would be significantly associated with weaker negative coupling between right dlPFC and right pHPC, but not aHPC. Finally, we examined whether any observed significant relationships were specific to TR-IMs by controlling for overall PTSD symptom severity.

## METHODS

### Participants

Trauma-exposed adults (N=136; 88 female) were recruited via advertisements in the local community and McLean Hospital outpatient programs. Study procedures were approved by the Mass General Brigham Human Research Committee, and all participants provided written informed consent. The main inclusion criteria were: 18 to 65 years of age; exposure to at least one DSM-5 Criterion A trauma for PTSD (1); and endorsement of at least two TR-IMs per week over the past month. Participants were also required to complete at least 70% of the daily surveys during the EMA period to be eligible. This threshold was based on precedent from the literature, including our prior publications in this and similar clinical samples (47). Detailed selection criteria can be found in the **Supplement**.

As previously described (45,46), participants completed a first study visit, a two-week EMA period, and a second visit, which included a clinical interview and MRI scanning. Twenty-seven participants were excluded due to: inability to tolerate the MRI scan (N=6), missing rsfMRI sequence (N=4), unusable rsfMRI due to excessive motion (N=13), poor structural-functional alignment (N=2), or structural brain abnormalities that could confound results (N=2). **Table 1** summarizes the characteristics of the final sample (N=109). Importantly, no significant differences were found in major demographic and clinical variables between our final sample (N=109) and participants excluded for completing less than 70% of EMA surveys (N=24) (see **Supplement**).

**Table 1.** Demographic and clinical characteristics of the sample of trauma-exposed adults (N = 109)

| **Characteristic** | **Mean (SD) or N (%)** |
| --- | --- |
| **Age** | 31.84 (9.89) |
| **Sex assigned at birth (female/male)** | 88 (80.73%) / 21 (19.27%) |
| **Race** |  |
| Asian | 5 (4.59%) |
| Bi-/multiracial | 20 (18.35%) |
| Black | 5 (4.59%) |
| White | 75 (68.81%) |
| Other | 3 (2.75%) |
| Unknown or Not Reported | 1 (0.92%) |
| **Hispanic Ethnicity** | 6 (5.50%) |
| **PTSD diagnosis/ subthreshold PTSD** | 85 (77.98%) / 24 (22.02%) |
| **CAPS-5 total score** | 34.32 (11.34) |
| **Total number of TR-IM surveys completed (out of 42)<sup>1</sup>** | 35.46 (3.81) |
| **TR-IM frequency** | 24.48 (26.11) |
| **TR-IM properties** |  |
| Intrusiveness | 2.71 (0.85) |
| Emotional intensity | 2.29 (0.82) |
| Vividness | 1.85 (0.91) |
| Visual properties | 2.51 (0.95) |
| Reliving | 1.64 (0.96) |

Note: Mean (standard deviation (SD)) is presented for continuous variables and number of participants (percentage) for categorical variables. <sup>1</sup>Only surveys assessing TR-IMs are included. CAPS-5: Clinician-Administered PTSD Scale for DSM-5; MINI: Mini International Neuropsychiatric Interview; PTSD: posttraumatic stress disorder; TR-IM: trauma-related intrusive memory.

### Clinical interviews

Doctoral-level clinicians administered the *Clinician-Administered PTSD Scale for DSM-5* (CAPS-5) (10) to assess whether participants met diagnostic criteria for PTSD and to determine overall symptom severity (i.e., CAPS-5 total score; **Table 1**). Additionally, the *Mini International Neuropsychiatric Interview* (M.I.N.I.) (48) version 7.0 was used to assess DSM-5 disorders, including those relevant to the exclusion criteria.

### Ecological Momentary Assessments (EMAs)

Our EMA protocol has been detailed elsewhere (46). Participants used the MetricWire smartphone application (MetricWire Inc., Kitchener, Ontario, Canada) to complete three daily surveys that queried TR-IM occurrence and properties over fourteen consecutive days (maximum 42 TR-IM surveys). TR-IM frequency equaled the total number of TR-IMs reported across all surveys during the two-week EMA period. TR-IM properties were assessed using a modified version of the Autobiographical Memory Questionnaire (49,50) and rated on a 0-4 Likert scale. For this report, we derived scores for intrusiveness, emotional intensity, vividness, visual properties, and reliving. Additional details on TR-IM frequency and properties are available in the **Supplement**.

### Imaging acquisition and preprocessing

Participants were scanned using a 3T Siemens MAGNETOM Prisma scanner (Siemens Healthineers, Erlangen, Germany) with a 64-channel head coil. The imaging protocol was adapted from the Human Connectome Project (https://www.humanconnectome.org/) and included a high-resolution MPRAGE T1-weighted sequence and a 13-minute eyes-open resting-state scan. The latter was preprocessed using fMRIPrep version 20.2.7 (51). Additional details are provided in the **Supplement**.

### Determination of regions of interest (ROIs)

Following a recent study that differentiated between aHPC and pHPC subregions to investigate right dlPFC-right HPC coupling during intrusion suppression in healthy and trauma-exposed individuals (13), we defined the right aHPC and pHPC ROIs as the right rostral and caudal hippocampus regions from the fine-grained connectivity-based Brainnetome atlas (52). The right dlPFC was defined as the combination of area 46 and ventral area 9/46 in the right middle frontal gyrus, based on the same atlas (52). Each ROI was eroded using a 2-mm Gaussian kernel to exclude inconsistent voxels at its boundaries (**Figure 1**). Using CONN toolbox version 22a (53), we performed two whole-brain seed-to-voxel resting-state functional connectivity analyses with the aHPC and pHPC as seeds to identify right dlPFC voxels showing significant positive and negative correlations with these subregions at the group level. This first step showed that the right dlPFC demonstrated both positive and negative coupling with the aHPC and pHPC (**Figures 2A** and **2B**, respectively). Because the focus of this study was on negative coupling between right dlPFC and HPC, we identified the right dlPFC voxel demonstrating the strongest anticorrelation with aHPC and the voxel demonstrating the strongest anticorrelation with pHPC for each participant. Four-millimeter spheres were centered around these participant-specific right dlPFC coordinates. These spheres were then re-masked using the suitable dlPFC mask to ensure that only voxels inside this ROI were included. This process yielded participant-specific right dlPFC ROIs anticorrelated with aHPC and pHPC (**Figures 3A** and **3B**, respectively).

**Figure 1.**
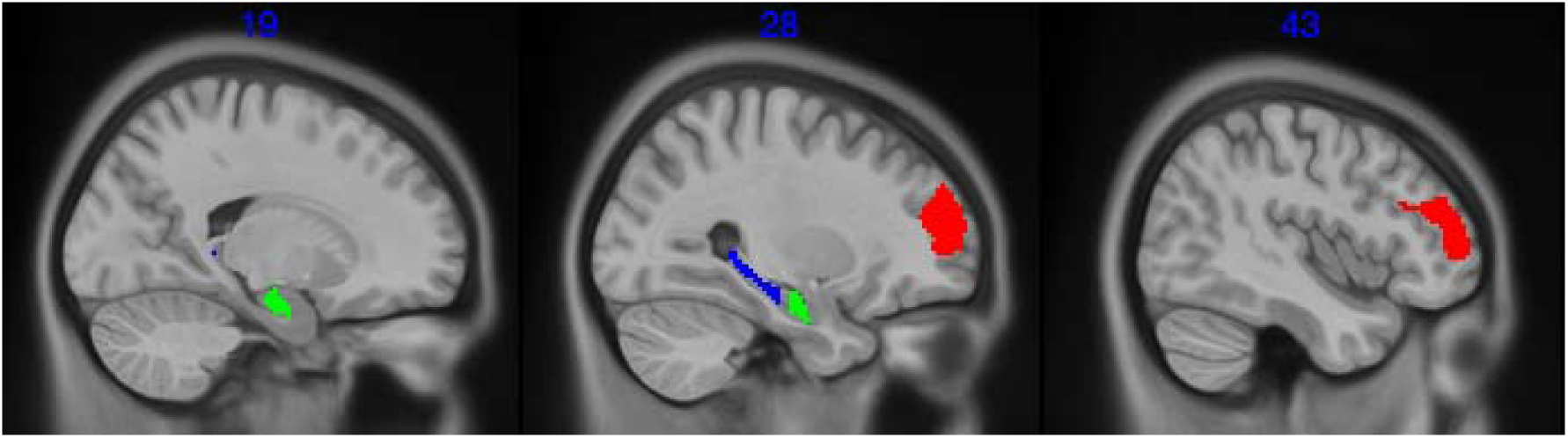
Regions of interest in MNI space. Using the Brainnetome atlas (52), we defined the right pHPC as the right caudal hippocampus (blue), the right aHPC as the right rostral hippocampus (green), and the right dlPFC as the combination of area 46 and ventral area 9/46 in the right middle frontal gyrus (red). Each region was eroded using a 2mm Gaussian kernel. aHPC: anterior hippocampus; dlPFC: dorsolateral prefrontal cortex; MNI: Montreal Neurological Institute; pHPC: posterior hippocampus.

**Figure 2.**
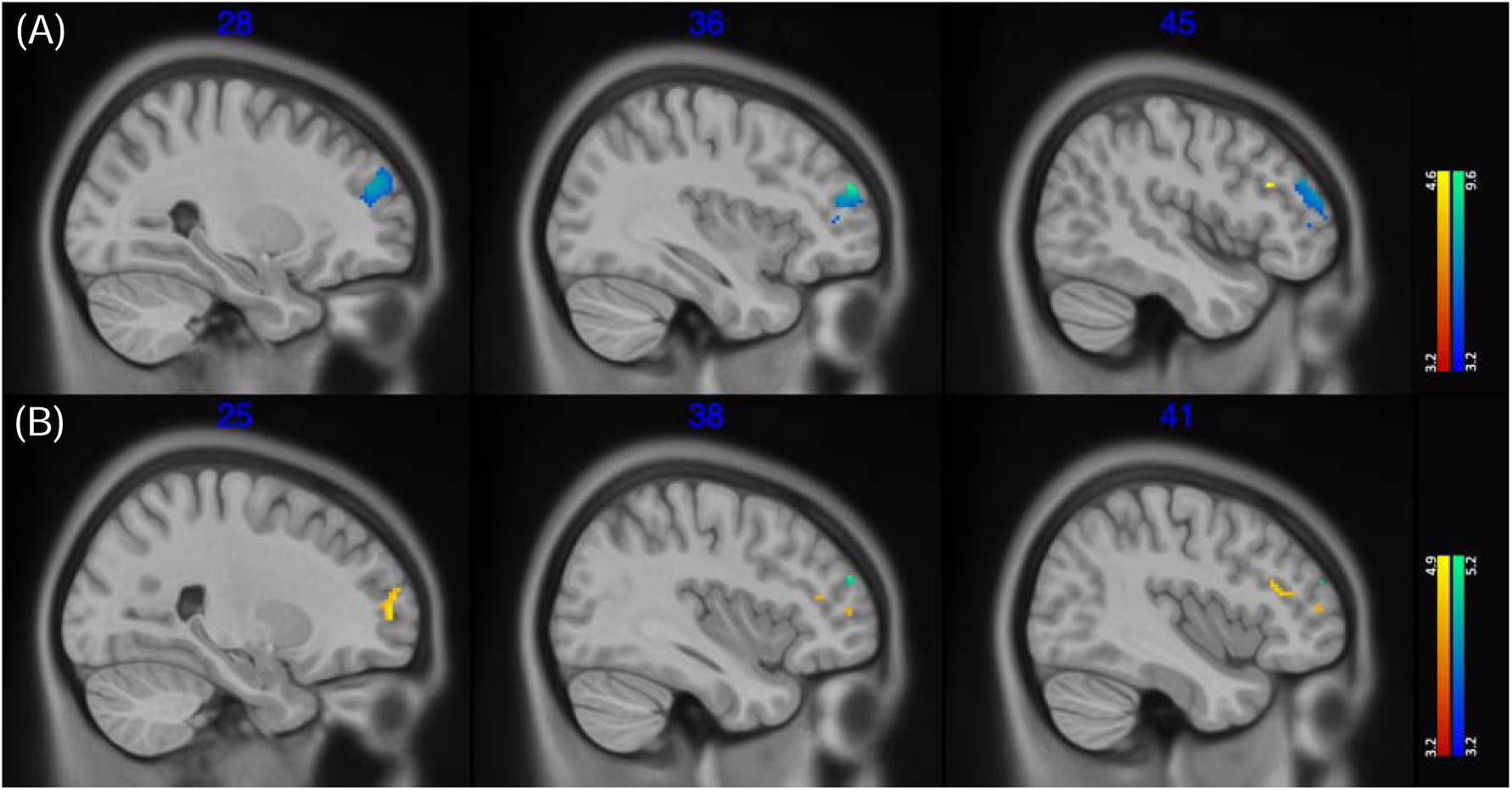
Seed-to-voxel functional connectivity results at the group level using either the aHPC (A) or pHPC (B) as seeds. Thresholded t-maps (voxel threshold: *p*-uncorrected < .001; cluster threshold: *p*_FDR_ < .05) showing voxels within the right dlPFC displaying significant positive (warm color) or negative (cold color) connectivity with aHPC (A) or pHPC (B) at the group level. aHPC: anterior hippocampus; dlPFC: dorsolateral prefrontal cortex; FDR: false discovery rate; MNI: Montreal Neurological Institute; pHPC: posterior hippocampus.

**Figure 3.**
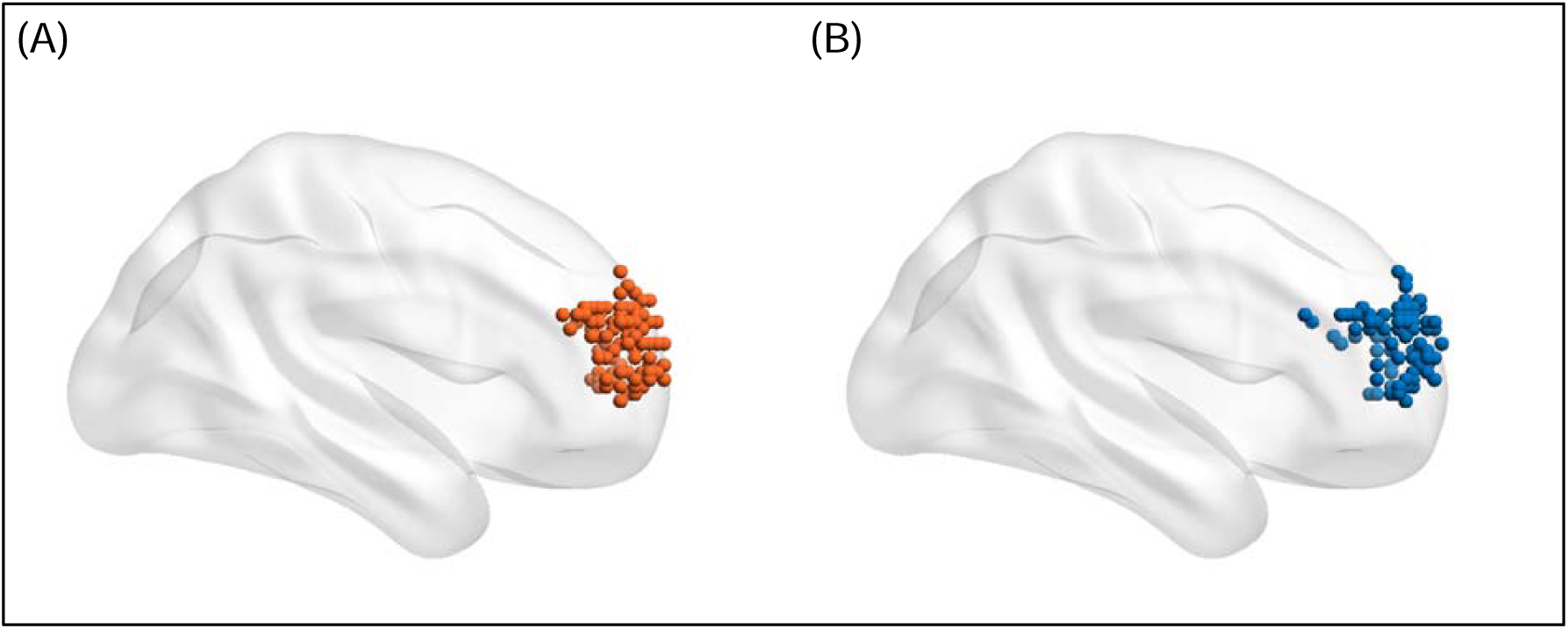
Participant-specific right dlPFC regions of interest. Dots represent the locations of the voxel within the right dlPFC demonstrating the strongest anticorrelation with either the right (A) anterior hippocampus or (B) posterior hippocampus for each participant. Dots are presented on a smoothed MNI152 surface (lateral view of right hemisphere) using BrainNet Viewer (69). dlPFC: dorsolateral prefrontal cortex; MNI: Montreal Neurological Institute.

### Resting-state functional connectivity analysis

An ROI-to-ROI analysis was performed using the right aHPC and pHPC atlas masks and the participant-specific right dlPFC ROIs using the CONN toolbox (53). Pairwise Fisher-transformed Pearson correlation coefficients of blood-oxygen-level-dependent time series were extracted for each participant. These individual transformed anticorrelations between the right dlPFC-right aHPC and right dlPFC-right pHPC were used as independent variables in the statistical analyses.

### Statistical analyses

To investigate the relationship between TR-IM frequency and connectivity patterns, we constructed two quasi-Poisson regression models. These models examined the association of TR-IM frequency, as the outcome variable, with the anticorrelation between the right dlPFC and the right aHPC or pHPC as predictor variables. Covariates in the models included age, sex, and total number of TR-IM surveys completed, with the duration of the EMA period (in days) included as an offset term. The small correlation between the total number of TR-IM surveys completed and duration of EMA period (r = −0.17; t = −1.831; *p* = .070) supported the inclusion of both variables in the quasi-Poisson models.

For the analysis of TR-IM properties, ten linear mixed-effects models (LMMs) were developed using the lme4 package (v1.1.34; (54)). These models evaluated the relationships of TR-IM properties (i.e., intrusiveness, emotional intensity, vividness, visual properties, and reliving) over time as outcomes, with the anticorrelation between the right dlPFC and the right aHPC (5 models) or pHPC (5 models) as predictors. Each model included a participant-specific random intercept. Age, sex, total number of TR-IM surveys completed, and EMA period duration were included as covariates. To verify that observed relationships of TR-IM frequency or properties with resting-state functional connectivity were not solely attributable to overall illness severity, CAPS-5 total score was added as a covariate in the models where significant findings were identified.

Correction for multiple comparisons was applied using the false discovery rate (FDR) method (55). The two-tailed significance threshold was set at *p_FDR_* < .05 for all analyses, and all predictors and covariates were centered and scaled. All statistical analyses were conducted using R software version 4.2.2 (56). Details of the *a priori* power and sample size calculations are provided in the **Supplement**.

## RESULTS

### Characterization of TR-IM frequency and properties during the EMA period

Participants completed an average of 35.46 surveys out of 42 (standard deviation (SD) = 3.81; range: 27-42), resulting in a data return rate of 84.43%. Over the two-week EMA period, they reported experiencing an average of 24.48 TR-IMs (SD = 26.11; range: 1-153; **Table 1**). TR-IM property scores were rated as follows: intrusiveness (M = 2.71, SD = 0.85; range: 0.25-4), emotional intensity (M = 2.29, SD = 0.82; range: 0.33-4), vividness (M = 1.85, SD = 0.91; range: 0-4), visual properties (M = 2.51, SD = 0.95; range: 0.20-4), and reliving (M = 1.64, SD = 0.96; range: 0-4).

### TR-IM frequency and right dlPFC-right aHPC/pHPC anticorrelation

The results of the two quasi-Poisson models are detailed in **Table S1**. They revealed no significant association between TR-IM frequency and right dlPFC-right aHPC anticorrelation (incidence rate ratio (IRR) = 1.07; t = 0.703; *p* = .482; *p_FDR_* = .578; 95% confidence interval (CI) = [0.88, 1.32]) and right dlPFC-right pHPC anticorrelation (IRR = 0.94; t = −0.566; *p* = .571; *p*_FDR_ = .623; 95% CI = [0.78, 1.15]).

### TR-IM properties in relation to right dlPFC-right aHPC anticorrelation

Results of the LMMs assessing relationships of TR-IM properties with right dlPFC-right aHPC anticorrelation are presented in **Table S2**. There was a significant positive association between right dlPFC-right aHPC anticorrelation and TR-IM emotional intensity (b = 0.20; t = 2.827; *p* = .005; *p_FDR_* = .044; 95% CI = [0.06, 0.34]) (**Figure 4A**). This association remained significant when controlling for CAPS-5 total score (b = 0.18; t = 2.671; *p* = .008; *p_FDR_* = .013; 95% CI = [0.05, 0.32]).

**Figure 4.**
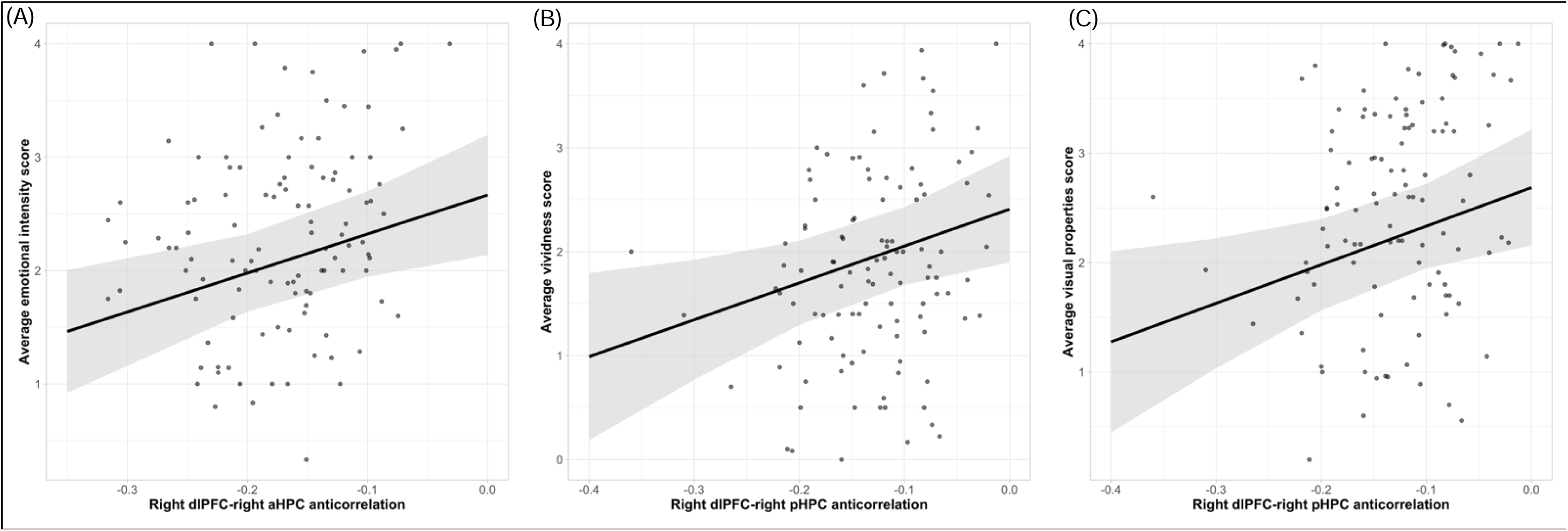
Relationships of TR-IM properties with right dlPFC-right aHPC and right dlPFC-pHPC anticorrelations. Scatterplots represent relationships of (A) TR-IM emotional intensity with right dlPFC-right aHPC anticorrelation, (B) TR-IM vividness with right dlPFC-right pHPC anticorrelation, and (C) TR-IM visual properties with right dlPFC-right pHPC anticorrelation. Dots represent raw data. Lines represent estimates of the fixed effect of interest from linear mixed-effects models, and light grey ribbons are 95% confidence intervals. aHPC: anterior hippocampus; dlPFC: dorsolateral prefrontal cortex; pHPC: posterior hippocampus; TR-IM: trauma-related intrusive memory.

There were no significant associations between right dlPHC-right aHPC anticorrelation and other TR-IM properties: intrusiveness (b = 0.09; t = 1.160; *p* = .246; *p_FDR_*= .422; 95% CI = [−0.06, 0.25]), vividness (b = 0.08; t = 1.033; *p* = .302; *p_FDR_* = .453; 95% CI = [−0.07, 0.24]), visual properties (b = 0.06; t = 0.739; *p* = .460; *p_FDR_* = .578; 95% CI = [−0.10, 0.22]), or reliving (b = 0.12; t = 1.466; *p* = .143; *p_FDR_*= .286; 95% CI = [−0.04, 0.29]; **Table S2**).

### TR-IM properties and right dlPFC-right pHPC anticorrelation

Results of the LMMs assessing relationships of TR-IM properties with right dlPFC-right pHPC anticorrelation are presented in **Table S3**. We found significant positive associations between right dlPFC-right pHPC anticorrelation and TR-IM vividness (b = 0.20; t = 2.547; *p* = .011; *p_FDR_* = .044; 95% CI = [0.05, 0.36]) and visual properties (b = 0.21; t = 2.550; *p* = .011; *p_FDR_* = .044; 95% CI = [0.05, 0.37]; **Figures 4B** and **4C**). These associations remained significant when controlling for CAPS-5 total score (vividness: b = 0.20; t = 2.492; *p* = .013; *p_FDR_* = .013; 95% CI = [0.04, 0.36]; visual properties: b = 0.21; t = 2.585; *p* = .010; *p_FDR_* = .013; 95% CI = [0.05, 0.37]).

There were no significant associations between right dlPFC-right pHPC and other TR-IM properties: intrusiveness (b = −0.00; t = −0.044; *p* = .965; *p_FDR_*= .965; 95% CI = [−0.16, 0.16]); emotional intensity (b = 0.11; t = 1.513; *p* = .130; *p_FDR_* = .286; 95% CI = [−0.03, 0.26]); reliving (b = 0.16; t = 1.858; *p* = .063; *p_FDR_* = .189; 95% CI = [−0.01, 0.33]; **Table S3**).

## DISCUSSION

This study investigated the relationships between TR-IM frequency and properties and resting-state functional connectivity between right aHPC/pHPC and anticorrelated right dlPFC regions in trauma-exposed individuals with PTSD symptoms. As hypothesized, TR-IM emotional intensity was positively associated with the connectivity of the right dlPFC with the right aHPC but not pHPC. Also as predicted, TR-IM vividness and visual properties were positively correlated with connectivity of the right dlPFC with the right pHPC but not aHPC. Contrary to expectations, no associations were found between TR-IM frequency, intrusiveness, or reliving and resting-state functional connectivity between the right dlPFC and either hippocampal subregion. These results suggest that certain phenomenological aspects of TR-IMs, but not their frequency, are linked to weaker intrinsic negative coupling between the right dlPFC and right aHPC/pHPC. Notably, all significant associations persisted after accounting for total PTSD symptom severity, indicating they are not merely a reflection of overall illness severity. These findings shed new light on the neural correlates of TR-IMs, highlighting the relevance of right dlPFC-right HPC functional negative coupling in understanding their emotional and sensory-perceptual properties.

Our findings indicate that right dlPFC-right HPC resting-state connectivity may primarily influence TR-IM sensory and emotional impact rather than their frequency. Indeed, stronger emotional intensity, greater vividness, and richer visual details of TR-IMs were associated with weaker right dlPFC-right HPC negative coupling at rest. In contrast, TR-IM frequency and intrusiveness were not significantly associated with resting-state functioning of this network. This suggests that intrinsic negative functional connectivity between these regions shapes only certain perceived properties of TR-IMs. Previous task-based fMRI studies have shown that voluntarily suppressing unwanted memories upregulates right dlPFC-right HPC functional negative coupling in healthy volunteers, correlating with fewer intrusions (13–18). This neural response appears to be impaired in PTSD, suggesting a disruption in the voluntary regulation of intrusive memories (13–15), though most of these studies did not specifically examine intrusion frequency. This earlier work also showed that engaging in top-down intrusion control can lead to sustained reductions in TR-IM occurrence and phenomenological properties, suggesting the involvement of tonically-operating control processes (57). These persistent changes may reflect plasticity within inhibitory circuitry, resulting in lasting increases in the functional inhibition of hippocampal-mediated memory retrieval. Our investigation of intrinsic connectivity within this circuit underscores this potential relevance of resting-state mechanisms in shaping the experience of TR-IMs, extending prior research demonstrating the importance of these intrinsic connectivity measures for task-related neural activity and behavior (24,25). These lines of evidence support the complementary roles of active and tonic intrusion control processes for the occurrence and phenomenological impact of TR-IMs.

Our findings also highlight the importance of distinguishing between anterior and posterior hippocampal-cortical networks when investigating TR-IM neurophysiology. As expected, TR-IM emotional intensity was significantly associated with right dlPFC-right aHPC, but not pHPC, negative coupling.

Conversely, TR-IM vividness and visual properties were significantly correlated with right dlPFC-right pHPC, but not aHPC, negative coupling. These findings align with previous research in healthy participants implicating the aHPC in the emotional processing of memories, and the pHPC in elaborating their sensory-perceptual details (30–37). In PTSD samples, only a few studies have examined resting-state functional connectivity of the aHPC/pHPC (45,58–60), with most focusing on overall PTSD symptom severity. In our earlier report on whole-brain spatiotemporal dynamics of the aHPC and pHPC in this sample, greater TR-IM emotional intensity was associated with fewer occurrences and less persistence of resting-state functional coactivation between the aHPC and areas of the medial prefrontal cortex and superior frontal gyrus (45). Combined with our current findings on negative coupling of the right dlPFC, this suggests that functional coordination of multiple prefrontal regions with the aHPC, rather than the pHPC, is relevant to TR-IM emotional intensity in PTSD. Furthermore, in our previous report, stronger TR-IM visual properties were associated with more occurrences of a coactivation pattern between HPC and visual and sensorimotor cortices and a deactivation pattern with precuneus, posterior cingulate cortex, angular gyrus, and superior frontal gyrus (45). Therefore, our findings indicate that dynamic and static negative functional coupling between HPC and multiple frontal regions may also contribute to stronger sensory-perceptual features of TR-IMs. However, due to methodological differences, the findings from both studies should be compared with caution. Indeed, based on *a priori* hypotheses, this study focused on a specific mechanism and circuit, namely the right dlPFC-right HPC static negative coupling. In contrast, our previous report employed a whole-brain dynamic approach that considered both positive and negative correlations (45). Nevertheless, these reports complement each other by highlighting the distinct roles of aHPC and pHPC and the interplay of activations/deactivations between prefrontal regions and HPC in TR-IM property neurophysiology in PTSD. These findings encourage consideration of functional differentiation within the HPC in future studies of task-based and resting-state functional correlates of trauma intrusion symptoms. By focusing on a specific circuit, the present study provides critical insights into potential neurotherapeutic targets to mitigate TR-IM emotional intensity and sensory vividness. While our previous report identified whole-brain network-wide relationships of aHPC and pHPC coactivation patterns, this study suggests a more precise target for potential non-invasive brain stimulation based on individual-level connectivity profiles (61). This offers a critical launching pad for future mechanistic and therapeutic studies addressing the pervasive and debilitating symptoms of TR-IMs.

This study has several strengths that increase confidence in the findings. First, using EMA to capture TR-IM frequency and properties minimizes the risk of recall bias associated with retrospective self-reports (62). Second, we accounted for individual differences in dlPFC function by defining participant-specific right dlPFC ROIs. This approach aligns with recent neuroimaging research advocating for individualized targets in neuromodulation interventions (61). Additionally, we separately considered anterior and posterior hippocampus subregions, given evidence of their distinct roles in autobiographical memory. Finally, our trauma-exposed sample represented a range of PTSD symptom severity, with approximately one quarter having subthreshold clinical presentations. This is critical because TR-IMs can cause clinically significant distress and impairment “in their own right” (63) and aligns with calls for improved understanding of the mechanisms of TR-IMs to inform more targeted treatments (6,64,65).

Limitations of our study also need to be acknowledged. While we considered a/pHPC subregions separately, emerging evidence suggests that hippocampal function may be better understood as a gradient, rather than as distinct anatomical divisions (28,66). Future investigations should incorporate gradient analyses to better capture the continuum of hippocampal function. Additionally, this study was not designed as a case-control study or to assess potential diagnostic group differences. Because TR-IMs also occur in other psychiatric disorders with high rates of trauma exposure, such as major depressive disorder (67), further research is needed to determine whether our findings are specific to PTSD or generalizable to other psychiatric conditions.

While this study indicates a role for right dlPFC-right HPC negative coupling at rest in TR-IM properties, it does not address how this resting-state coupling might relate to active increases in negative coupling during voluntary intrusion suppression. Previous task-based fMRI studies have not explored this connection, and further research is required to understand how resting-state functional connectivity relates to active brain mechanisms underlying TR-IMs (13–15). Additionally, the present study cannot determine whether weaker right dlPFC-right aHPC/pHPC functional negative coupling at rest represents a premorbid risk factor for more intense TR-IMs, a consequence of trauma exposure, or a combination of both. Given evidence of structural plasticity in the dlPFC after trauma exposure (68), future longitudinal studies, particularly in the immediate aftermath of trauma, should investigate whether intrinsic right dlPFC-right HPC negative coupling predicts more vivid, emotionally intense, and visually detailed TR-IMs. Finally, the present study focused on static functional connectivity, preventing us from drawing conclusions regarding the directionality of the identified functional patterns. Future studies using dynamic techniques are warranted to examine relationships between TR-IM properties and effective resting-state connectivity of the right dlPFC-right HPC circuit.

## Conclusion

This study provides novel insights into the neural mechanisms of TR-IMs, highlighting the relevance of right dlPFC-right HPC resting-state negative coupling as a potential biomarker of their emotional impact and sensory-perceptual properties. Our findings also reinforce the importance of differentiating the roles of the anterior and posterior hippocampi in trauma memory processes. These findings build on literature implicating phasic engagement of this circuit during voluntary attempts to suppress intrusive memories. Altogether, it seems reasonable to posit that phasic and tonic inhibitory mechanisms mediated by right dlPFC-right HPC circuits may be differentially relevant to component processes of intrusive reexperiencing in PTSD. Their complementary roles deserve further investigation, given the urgent need to address one of the most debilitating and treatment-refractory symptoms of trauma in an increasingly traumatic world.

## Data Availability

Data can be obtained from the corresponding author (IMR) upon completion of a data sharing agreement.

## ACKNOWLEDGMENTS

<u>Funding</u>: This research was supported by NIMH award R01-MH120400 (IMR). Additionally, IMR was partially supported by NIMH award P50-MH115874 (Project 4 PIs: IMR, Scott L. Rauch; PDs: William A. Carlezon Jr. and Kerry J. Ressler). <u>Author contributions (based on contributor roles taxonomy)</u>: Conceptualization: QD, KJC, IMR; Methodology: QD, KJC, BR, JTB, IMR; Software: QD, BR, JTB, YP; Validation: QD, KJC, BR; Formal analysis: QD, BR; Investigation: KJC, YP, IMR; Resources: IMR; Data curation: QD, KJC, BR, YP, IMR; Visualization: QD, BR; Supervision: BR, IMR; Project administration: IMR; Funding acquisition: IMR; Writing - original draft: QD, IMR; Writing - review and editing: all authors. <u>Data availability</u>: Data can be obtained from the corresponding author (IMR) upon completion of a data sharing agreement.

## SUPPLEMENT

### METHODS

#### Participants

The inclusion criteria were: (a) 18 to 65 years of age; (b) exposure to at least one DSM-5 Criterion A trauma for posttraumatic stress disorder (PTSD) (1); (c) endorsement of at least two trauma-related intrusive memories (TR-IM)s per week in the past week; (d) sufficient proficiency in English to complete study procedures; (e) access to a smartphone compatible with the MetricWire application (MetricWire Inc., Kitchener, Ontario, Canada). In addition, the exclusion criteria were: (a) left-handedness; (b) medical condition that could confound results (e.g., seizure disorder); (c) current psychotic disorder or manic mood episode; (d) history of moderate-to-severe traumatic brain injury, or head trauma with loss of consciousness > 5 minutes; (e) past month moderate-to-severe alcohol or substance use disorder; (f) contraindications for magnetic resonance imaging (MRI); (g) positive pregnancy test for female participants on the day of MRI scanning; (h) report of experiencing intrusions only as thoughts, not as memories; (i) completion of less than 70% of the daily surveys (i.e., < 49 surveys) during the ecological momentary assessment (EMA) period.

When comparing our final sample (N = 109) to participants excluded from the study (N = 24) due to completing less than 70% of daily surveys, no significant differences were found in terms of age (*p* = .207), overall PTSD symptom severity measured by the PTSD Checklist for DSM-5 (PCL-5) total score at baseline (*p* = .393) (2), number of TR-IMs per day (*p* = .940), or average TR-IM properties over the EMA period (all p’s > .05) using unpaired two-samples Wilcoxon rank sum test. There also was no significant group difference in sex distribution (*p* = .999) using Fisher’s exact test.

#### Ecological Momentary Assessment (EMA)

TR-IM occurrences were queried at the beginning of each TR-IM survey (“*Since the last time you completed a survey, did you have any unwanted memories of your trauma? (If “Yes”: How many?”)*. If a participant reported having experienced TR-IMs since the last completed survey, further questions on the phenomenological properties associated with these TR-IMs were asked using an adapted version of the Autobiographical Memory Questionnaire (AMQ) (3,4). In the present study, we derived five TR-IM property scores from the AMQ (3,4): (a) intrusiveness (“*This memory came to me out of the blue, without my trying to think about it”*); (b) emotional intensity (“*As I was remembering the event, the emotions I felt were intense*”); (c) vividness (i.e., average score across the following two items: “*My memory of this event had lots of details*”, and “*My memory of this event was clear, not fuzzy or cloudy*”); (d) visual properties (i.e., average score across the following five items: “*As I was remembering the event, I could see it in my mind*”, “*As I was remembering the event, I could identify or name the setting where the memory occurred*”, “*As I was remembering the event, I experienced a scene in which elements of the setting were located relative to each other*”, “*As I was remembering the event, I could identify the people and things that were in the memory*”, and “*As I was remembering the event, I could describe where the people and things were located in the memory*”); and (e) reliving (i.e., average score across the following two items: “*As I was remembering the event, it was as if I was reliving it*”, and “*As I was remembering the event, it was as if I traveled back to the time it happened*”).

#### Imaging acquisition

Structural sequences were initially acquired using the Human Connectome Project (HCP; https://www.humanconnectome.org/) Young Adult protocol (N = 7). Early in the study, the imaging protocol was transitioned to the HCP Lifespan protocol (N = 102). These two protocols were developed to be compatible (5). The imaging protocol included: (a) an anatomical high-resolution MPRAGE T1-weighted sequence (parameters for the HCP Young Adult protocol: voxel size = 0.7 mm isotropic; TR = 2,400 ms; TE = 2.14 ms; FOV = 224 x 210 mm; matrix size = 208 x 300 x 320 slices; flip angle = 8°; parameters for the HCP Lifespan protocol: voxel size = 0.8 mm isotropic; TR = 2,500 ms; TEs = 1.81/3.6/5.39/7.27 ms; FOV = 256 x 240 mm; matrix size = 208 x 300 x 320 slices; flip angle = 8°), and (b) an eyes-open resting-state T2-weighted echoplanar images (parameters for the HCP Young Adult protocol: TR = 720; TE = 33 ms; in-plane resolution = 2 mm; voxel size = 2 mm isotropic; multiband factor = 8; anterior-posterior phase encoding = one run of 1200 frames, ∼13 minutes in length; parameters for the HCP Lifespan protocol: TR = 800; TE = 37 ms; in-plane resolution = 2 mm; voxel size = 2 mm isotropic; multiband factor = 8; anterior-posterior phase encoding = one run of 976 frames, ∼13 minutes in length). Each image was visually inspected to ensure the absence of artifacts and anatomical abnormalities before inclusion in the analyses, resulting in the exclusion of two participants.

#### Resting-state functional MRI data preprocessing

Resting-state fMRI data were preprocessed using fMRIPrep version 20.2.7 (6). T1-weighted images were corrected for intensity non-uniformity (7). Brain tissue segmentation of cerebrospinal fluid (CSF), white matter (WM), and gray matter (GM) was performed on the brain-extracted T1-weighted (8). Brain surfaces were reconstructed using recon-all (9), and the brain mask estimated previously was refined with a custom variation of the method to reconcile Advanced Normalization Tools (ANTs)-derived and FreeSurfer-derived segmentations of the cortical GM of Mindboggle (10). Volume-based spatial normalization to Montreal Neurological Institute (MNI) standard space (MNI152NLin6Asym) was performed through nonlinear registration with antsRegistration (ANTs version 2.3.3), using brain-extracted versions of both T1-weighted reference and the T1-weighted template.

Echo-planar images (EPI) were corrected for susceptibility distortions using the fMRIPrep fieldmap-less approach (11). Based on the estimated susceptibility distortion, a corrected EPI reference was calculated for a more accurate co-registration with the anatomical reference. The reference was co-registered to the T1-weighted reference with six degrees of freedom (12). Head motion parameters with respect to the reference (transformation matrices and six corresponding rotation and translation parameters) were estimated before any spatiotemporal filtering (13). EPI images were slice-time corrected (14). The time series were resampled onto their original, native space by applying a single composite transform to correct for head motion and susceptibility distortions. The time series were resampled into standard space, generating a preprocessed run in MNI152NLin6Asym space. First, a reference volume and its skull-stripped version were generated using a custom methodology of fMRIPrep. Automatic removal of motion artifacts using independent component analysis (ICA-AROMA) (15) was performed on the preprocessed images on MNI space time-series after removal of non-steady state volumes and spatial smoothing with an isotropic, Gaussian kernel of 6mm FWHM (full-width half-maximum). Corresponding “non-aggressively” denoised runs were produced after such smoothing (15).

Additional preprocessing was conducted using the CONN toolbox version 22a (16) with Statistical Parametric Mapping version 12 (7771) (SPM12; Wellcome Department of Cognitive Neurology, London, UK; www.fil.ion.ucl.ac.uk/spm) within Matlab version R2023a (MathWorks, Natick, MA, USA) and included the regression of physiological noise from white matter and cerebrospinal fluid using the CompCor method (17), scrubbing of motion outliers (framewise displacement (FD) > 0.5 mm) (18), and high pass (0.01 Hz) filtering. Participants were excluded if their total mean FD across volumes exceeded 0.5 mm or greater than 20% of volumes exceeded FD ≥0.5 mm (N = 13) (18).

#### Power and sample size calculations

To estimate the sample size needed for two two-tailed Poisson regression models, we used WebPower R package (https://cran.r-project.org/web/packages/WebPower/WebPower.pdf) with the following parameters:

– Significance level *α* corrected using the false discovery rate (FDR) method of 0.0375 (computed as the mean FDR *α* for *m* tests using 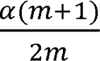, where m = 2 and *α* = .05) (19)
– Base rate of 1 (i.e., one TR-IM per day)
– Effect sizes (i.e., incidence rate ratio (IRR)) varying from 1.68 (small) to 3.47 (medium) to 6.71 (large)) (20)
– Power of 80%

The plot below represents the minimum sample size as a function of the effect size with the abovementioned parameters. The dashed red line represents the sample size of the present study (N = 109). This plot illustrates that the sample size of the present study (N = 109) was sufficient to detect small to medium effect sizes with a power of 80%.

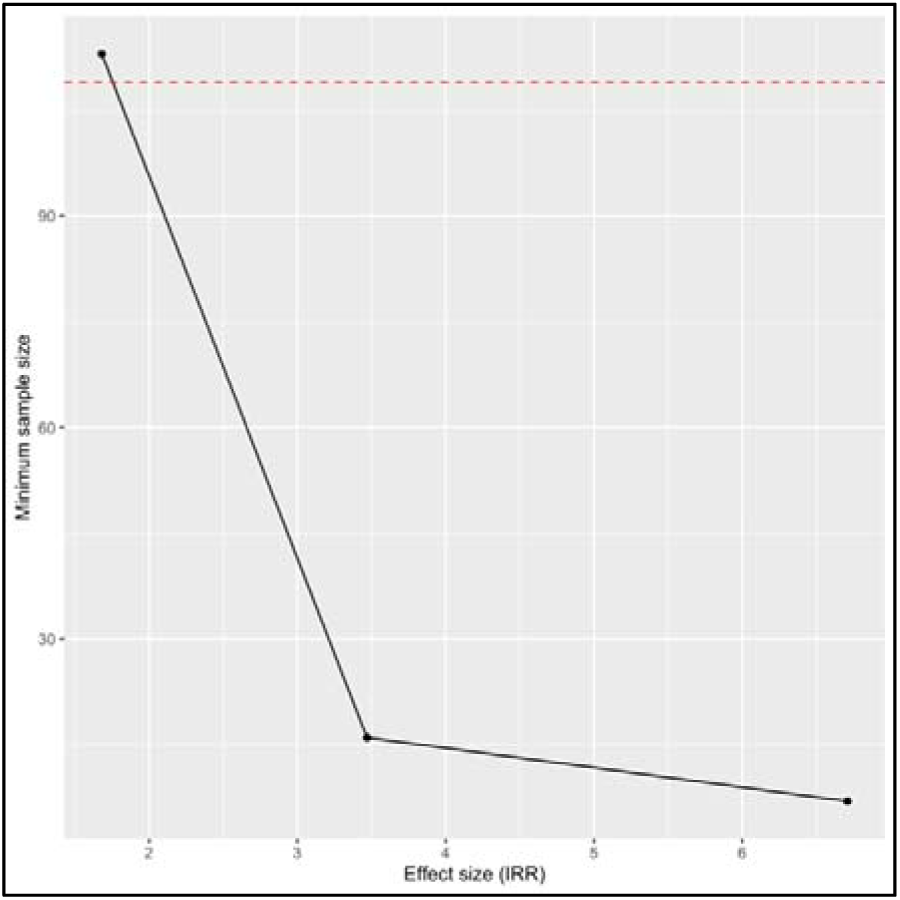

Power calculations for linear mixed-effects models require simulating data based on parameters derived from the experience or literature. However, because there is no precedence in the literature, we could not estimate reliably within what range our predictors of interest would fall, making data simulation challenging. Therefore, we performed power calculations for linear models instead. We used the WebPower R package (https://cran.r-project.org/web/packages/WebPower/WebPower.pdf) to estimate the sample size needed for ten two-tailed linear models with the following parameters:

– Significance level FDR-corrected *α* of 0.0275 (computed as the mean *α* for *m* tests using, where m = 10 and *α* = .05) (19)
– Effect sizes f varying from 0.02 (small) to 0.15 (medium) to 0.25 (large)) (21)
– Number of predictors in full model p1 = 4
– Number of predictors in reduced model p2 = 3
– Power of 80%

The plot below represents the minimum sample size as a function of the effect size with the abovementioned parameters. The dashed red line represents the sample size of the present study (N = 109). This plot illustrates that the sample size of the present study (N = 109) was sufficient to detect small to medium effect sizes with a power of 80%.

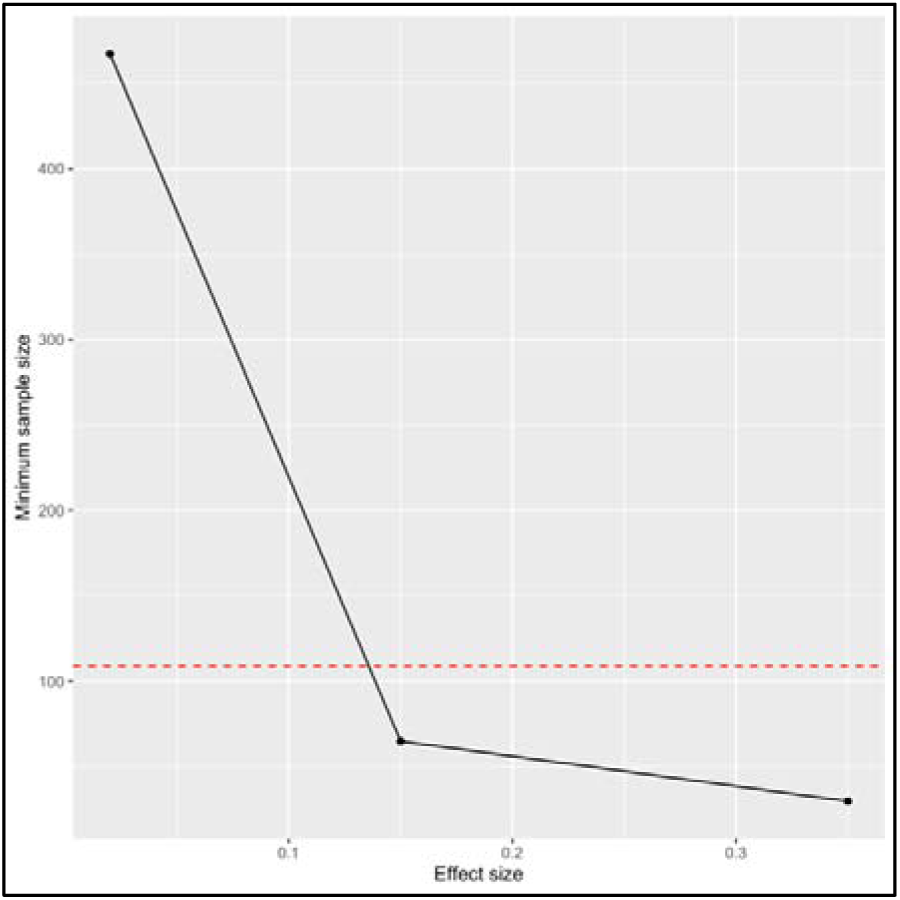

---

**Table S1.** Results of quasi-Poisson regression models assessing the relationships of TR-IM frequency with right dlPFC-right aHPC (top) and pHPC (bottom) resting-state anticorrelation.

|  | TR-IM frequency |  |  |  |
| --- | --- | --- | --- | --- |
|  | IRR | t-value | p-value/<br>p<sub>FDR</sub>-value | 95% CI |
| Right dlPFC-right aHPC anticorrelation | 1.07 | 0.703 | .482/<br>.578 | [0.88, 1.32] |
| Age | 1.08 | 0.848 | .397 | [0.89, 1.32] |
| Sex | 0.88 | -0.526 | .599 | [0.54, 1.48] |
| Total number of TR-IM surveys completed | 1.05 | 0.466 | .641 | [0.86, 1.28] |
| **Nagelkerke R<sup>2</sup>** | 0.399 |  |  |  |
|  | TR-IM frequency |  |  |  |
| Right dlPFC-right pHPC anticorrelation | 0.94 | -0.566 | .571/<br>.623 | [0.78, 1.15] |
| Age | 1.09 | 0.366 | .364 | [0.90, 1.32] |
| Sex | 0.89 | -0.490 | .624 | [0.55, 1.48] |
| Total number of TR-IM surveys completed | 1.06 | 0.551 | .582 | [0.87, 1.30] |
| **Nagelkerke R<sup>2</sup>** | 0.369 |  |  |  |

Note: Only *p*-values associated with the predictors of interest (i.e., right dlPFC-right aHPC or pHPC anticorrelation) are corrected using the False Discovery Rate (FDR) method (19). aHPC: anterior hippocampus; CI: confidence interval; dlPFC: dorsolateral prefrontal cortex; FDR: false discovery rate; IRR: incidence rate ratio; pHPC: posterior hippocampus; TR-IM: trauma-related intrusive memory.

**Table S2.**
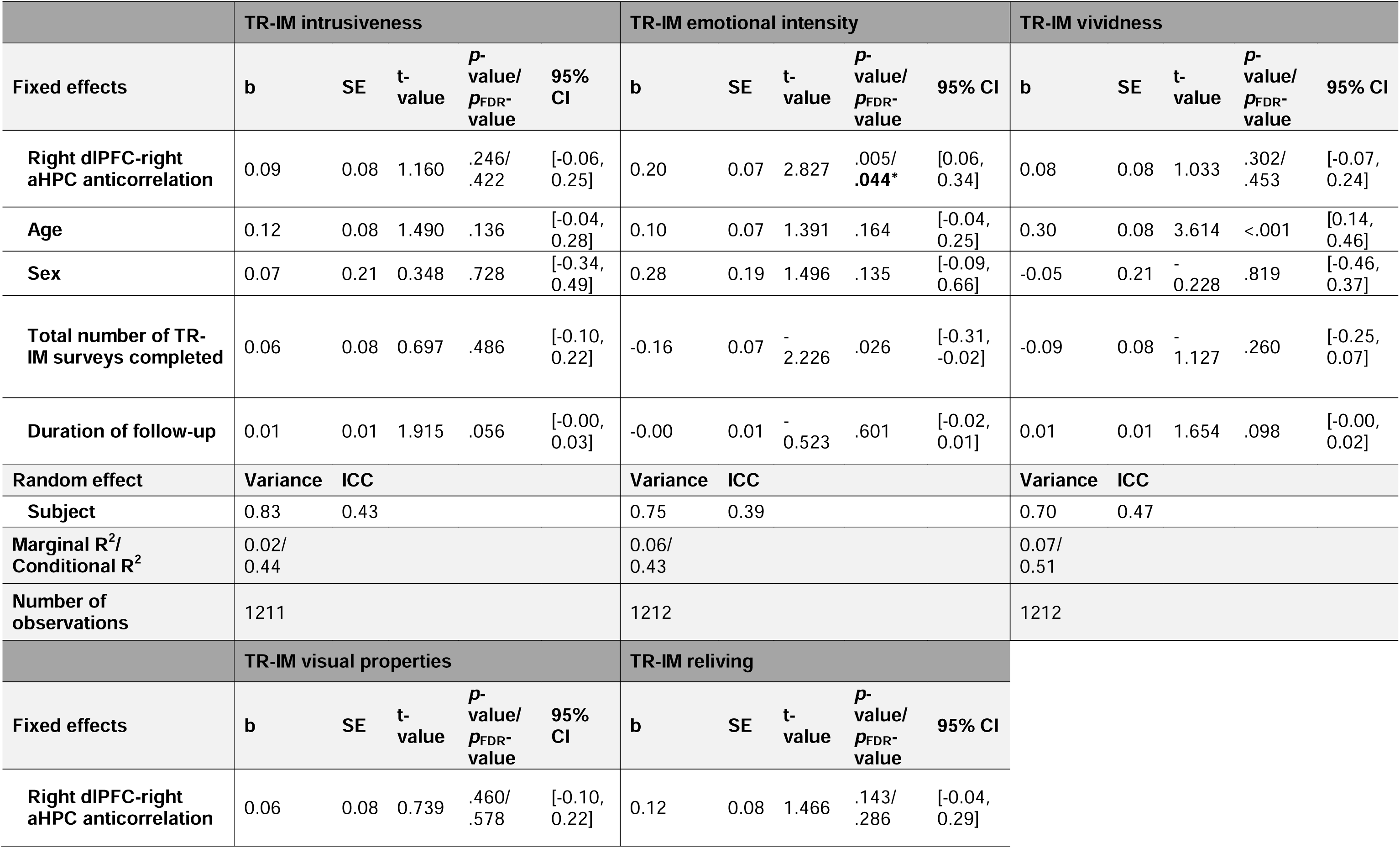
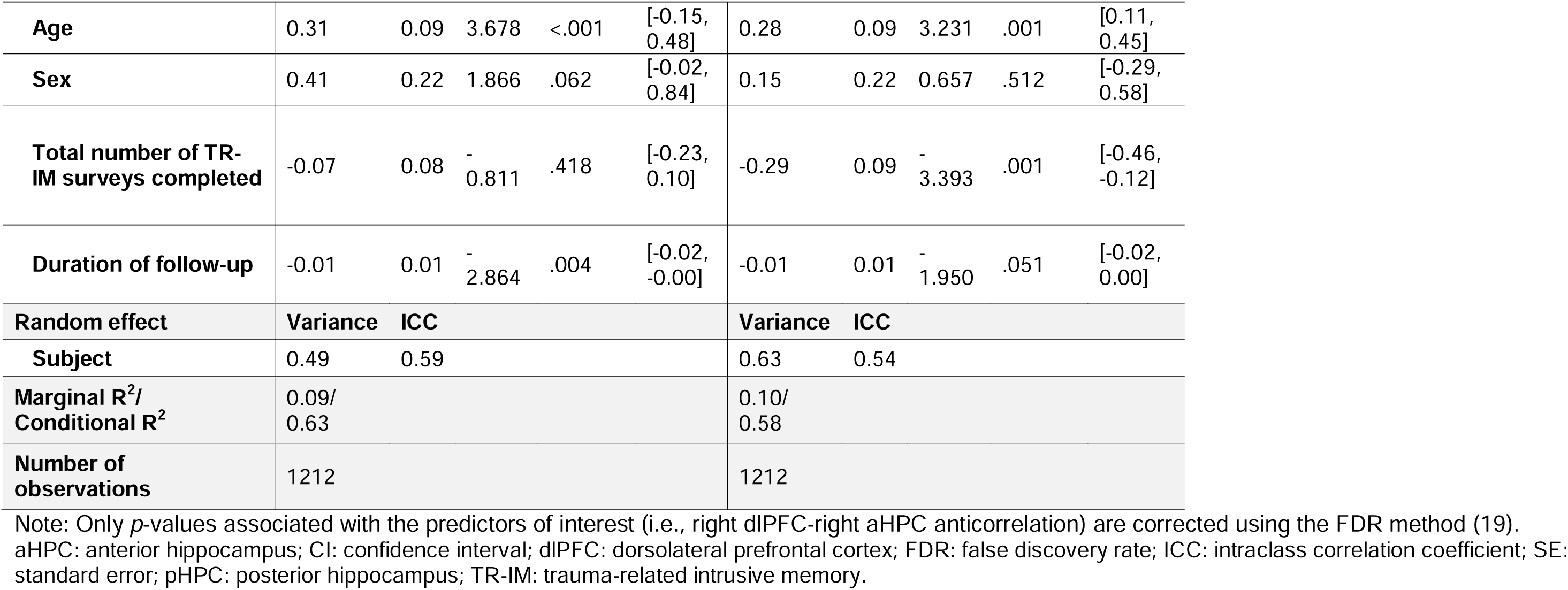
Results of linear mixed-effects models assessing the relationships of TR-IM properties with right dlPFC-right aHPC resting-state anticorrelation.

**Table S3.**
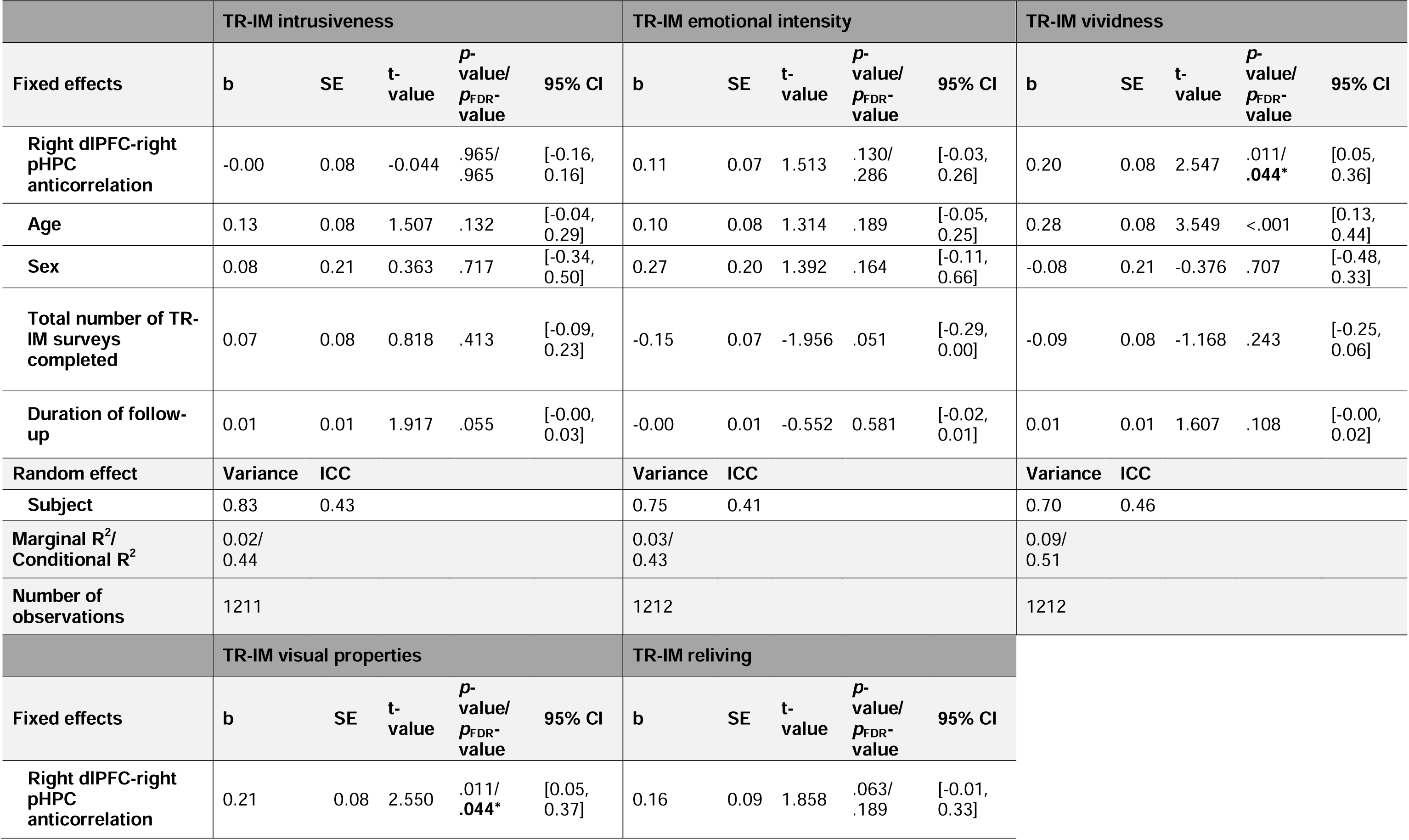
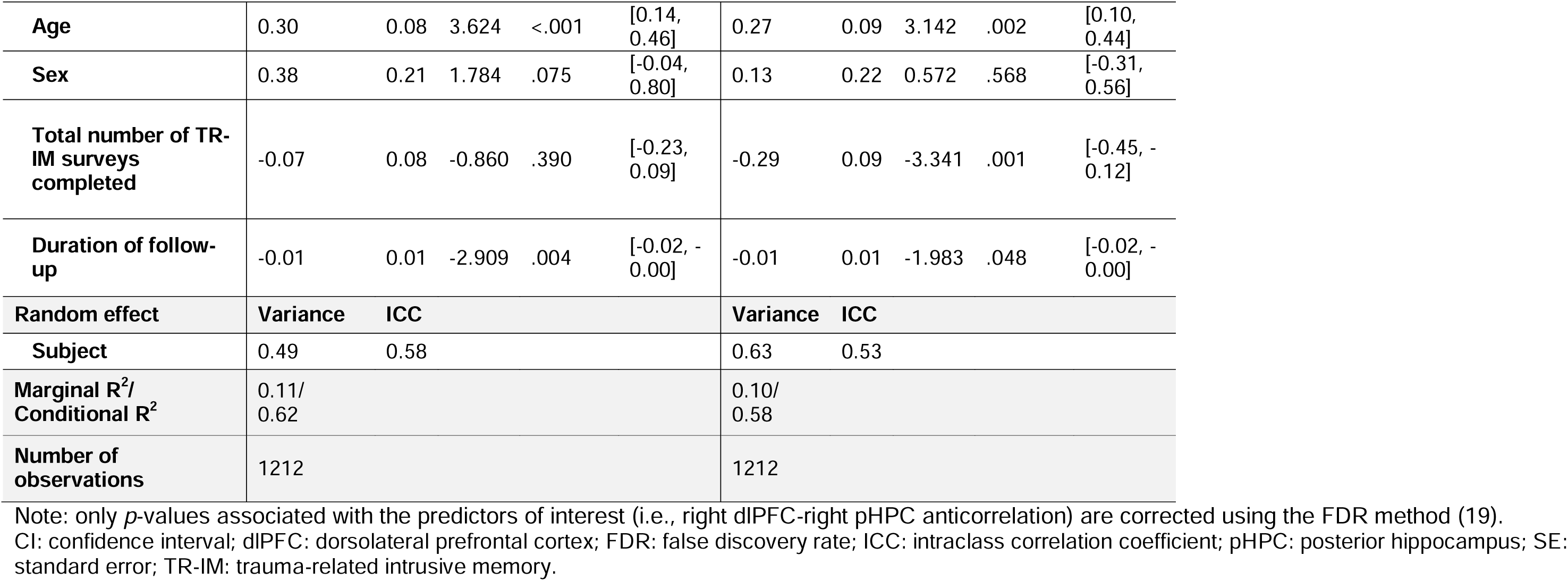
Results of linear mixed-effects models assessing the relationships of TR-IM properties with right dlPFC-right pHPC resting-state anticorrelation.

|  | TR-IM intrusiveness |  |  |  |  | TR-IM emotional intensity |  |  |  |  | TR-IM vividness |  |  |  |  |
| --- | --- | --- | --- | --- | --- | --- | --- | --- | --- | --- | --- | --- | --- | --- | --- |
| Fixed effects | b | SE | t-value | p-value/<br>p<sub>FDR</sub>-value | 95% CI | b | SE | t-value | p-value/<br>p<sub>FDR</sub>-value | 95% CI | b | SE | t-value | p-value/<br>p<sub>FDR</sub>-value | 95% CI |
| Right dlPFC-right pHPC anticorrelation | -0.00 | 0.08 | -0.044 | .965/.965 | [-0.16, 0.16] | 0.11 | 0.07 | 1.513 | .130/.286 | [-0.03, 0.26] | 0.20 | 0.08 | 2.547 | .011/.044* | [0.05, 0.36] |
| Age | 0.13 | 0.08 | 1.507 | .132 | [-0.04, 0.29] | 0.10 | 0.08 | 1.314 | .189 | [-0.05, 0.25] | 0.28 | 0.08 | 3.549 | <.001 | [0.13, 0.44] |
| Sex | 0.08 | 0.21 | 0.363 | .717 | [-0.34, 0.50] | 0.27 | 0.20 | 1.392 | .164 | [-0.11, 0.66] | -0.08 | 0.21 | -0.376 | .707 | [-0.48, 0.33] |
| Total number of TR-IM surveys completed | 0.07 | 0.08 | 0.818 | .413 | [-0.09, 0.23] | -0.15 | 0.07 | -1.956 | .051 | [-0.29, 0.00] | -0.09 | 0.08 | -1.168 | .243 | [-0.25, 0.06] |
| Duration of follow-up | 0.01 | 0.01 | 1.917 | .055 | [-0.00, 0.03] | -0.00 | 0.01 | -0.552 | 0.581 | [-0.02, 0.01] | 0.01 | 0.01 | 1.607 | .108 | [-0.00, 0.02] |
| Random effect | Variance | ICC |  |  |  | Variance | ICC |  |  |  | Variance | ICC |  |  |  |
| Subject | 0.83 | 0.43 |  |  |  | 0.75 | 0.41 |  |  |  | 0.70 | 0.46 |  |  |  |
| Marginal R<sup>2</sup>/Conditional R<sup>2</sup> | 0.02/0.44 |  |  |  |  | 0.03/0.43 |  |  |  |  | 0.09/0.51 |  |  |  |  |
| Number of observations | 1211 |  |  |  |  | 1212 |  |  |  |  | 1212 |  |  |  |  |
|  | TR-IM visual properties |  |  |  |  | TR-IM reliving |  |  |  |  |  |  |  |  |  |
| Fixed effects | b | SE | t-value | p-value/<br>p<sub>FDR</sub>-value | 95% CI | b | SE | t-value | p-value/<br>p<sub>FDR</sub>-value | 95% CI |  |  |  |  |  |
| Right dlPFC-right pHPC anticorrelation | 0.21 | 0.08 | 2.550 | .011/.044* | [0.05, 0.37] | 0.16 | 0.09 | 1.858 | .063/.189 | [-0.01, 0.33] |  |  |  |  |  |

|  |  |  |  |  |  |  |  |  |  |  |
| --- | --- | --- | --- | --- | --- | --- | --- | --- | --- | --- |
| **Age** | 0.30 | 0.08 | 3.624 | <.001 | [0.14, 0.46] | 0.27 | 0.09 | 3.142 | .002 | [0.10, 0.44] |
| **Sex** | 0.38 | 0.21 | 1.784 | .075 | [-0.04, 0.80] | 0.13 | 0.22 | 0.572 | .568 | [-0.31, 0.56] |
| **Total number of TR-IM surveys completed** | -0.07 | 0.08 | -0.860 | .390 | [-0.23, 0.09] | -0.29 | 0.09 | -3.341 | .001 | [-0.45, -0.12] |
| **Duration of follow-up** | -0.01 | 0.01 | -2.909 | .004 | [-0.02, -0.00] | -0.01 | 0.01 | -1.983 | .048 | [-0.02, -0.00] |
| **Random effect** | **Variance** | **ICC** |  |  |  | **Variance** | **ICC** |  |  |  |
| **Subject** | 0.49 | 0.58 |  |  |  | 0.63 | 0.53 |  |  |  |
| **Marginal R<sup>2</sup>/Conditional R<sup>2</sup>** | 0.11/<br>0.62 |  |  |  |  | 0.10/<br>0.58 |  |  |  |  |
| **Number of observations** | 1212 |  |  |  |  | 1212 |  |  |  |  |

Note: only *p*-values associated with the predictors of interest (i.e., right dlPFC-right pHPC anticorrelation) are corrected using the FDR method (19). CI: confidence interval; dlPFC: dorsolateral prefrontal cortex; FDR: false discovery rate; ICC: intraclass correlation coefficient; pHPC: posterior hippocampus; SE: standard error; TR-IM: trauma-related intrusive memory.

